# Automated Identification of Thrombectomy Amenable Vessel Occlusion on Computed Tomography Angiography using Deep Learning

**DOI:** 10.1101/2024.05.07.24306974

**Authors:** Jung Hoon Han, Hoyeon Lee, Gi-Hoon Park, Hotak Hong, Dongmin Kim, Jae Guk Kim, Joon-Tae Kim, Leonard Sunwoo, Chi Kyung Kim, Wi-Sun Ryu

**Author notes:** Correspondence: Chi Kyung Kim, MD, PhD, Department of Neurology, Korea University Guro Hospital and Korea University College of Medicine, 148 Gurodong-ro, Guro-gu, Seoul 08308, Korea,; Wi-Sun Ryu, MD, PhD, Research Center for Artificial Intelligence, JLK Inc., Seoul, 06141, Republic of Korea / /. Equally contributed as corresponding authors (Chi Kyung Kim, Wi-Sun Ryu).

## Abstract

**Objectives:** Recent advancements have extended the treatment window for large vessel occlusion in acute ischemic stroke, prompting a shift in the standard of care for patients presenting within 6 to 24 hours. We developed and externally validated an automated deep learning algorithm for detecting thrombectomy amenable vessel occlusion (TAVO) in computed tomography angiography (CTA).

**Methods:** The algorithm was trained on 2,045 acute ischemic stroke patients who underwent CTA, and validation was conducted using two external datasets comprising 64 (external 1) and 313 (external 2) patients with ischemic stroke. TAVO was defined as occlusion in the intracranial internal carotid artery (ICA), or M1/M2 segment of the middle cerebral artery (MCA). Utilizing U-Net for vessel segmentation and EfficientNetV2 for TAVO prediction, the algorithm’s diagnostic performance was assessed using the area under the receiver operating characteristics curve (AUC), sensitivity, specificity, positive predictive value (PPV), and negative predictive value (NPV).

**Results:** The mean age in the training and validation dataset was 68.7±12.6; 56.3% were men, and 18.0% had TAVO. The algorithm achieved AUC of 0.950 (95% CI, 0.915–0.971) in the internal test. For the external datasets 1 and 2, the AUCs were 0.970 (0.897–0.997) and 0.971 (0.924–0.990), respectively. Notably, the algorithm demonstrated robust sensitivity and specificity (approximately 0.95) for intracranial ICA or M1-MCA occlusion, but a slight reduction in performance for isolated M2-MCA occlusion.

**Conclusion:** This validated algorithm has potential applications in identifying TAVO and could aid less-experienced clinicians, potentially expediting the treatment process for eligible patients.

## Introduction

Advancements in stroke imaging and procedural devices have extended the endovascular treatment (EVT) window for patients with hyperacute ischemic stroke.^1,2^ The DAWN (DWI or CTP Assessment with Clinical Mismatch in the Triage of Wake-Up and Late Presenting Strokes Undergoing Neurointervention with Trevo) and DFFUSE 3 (Endovascular Therapy Following Imaging Evaluation for Ischemic Stroke) trials have changed the standard of care for ischemic stroke patients who present within 6 to 24 hours of their last well known status.

Triage entry for clinical trials primarily relies on magnetic resonance imaging (MRI) or computed tomography perfusion (CTP) to identify clinical or tissue mismatch and is now endorsed in guidelines. However, access to acute MRI or CTP is limited and not commonly available in the majority of primary stroke centers worldwide. Recent studies have brought attention to more readily available imaging techniques, such as CT angiography.^3^ The CT for Late Endovascular Reperfusion (CLEAR) trial^4^ revealed no significant differences in clinical outcomes between patients selected using non-contrast CT with CT angiography and those selected using CTP or MRI. In addition, a sub-study^5^ of the HERMES collaboration (Highly Effective Reperfusion Evaluated in Multiple Endovascular Stroke Trials) has extended this approach to the early time window (0-6 hours) by demonstrating that the rates of favorable functional outcomes were comparable between patients who underwent CTP and those who did not.

Two-thirds of EVT candidates were initially routed to centers not equipped for EVT,^6^ despite better outcomes and higher chances of receiving EVT at EVT-capable centers. Consequently, non-EVT-capable centers must consistently identify large vessel occlusion (LVO) around the clock, ensuring quick reporting to facilitate patient transfer to EVT-capable centers. However, a lack of vascular specialists poses challenges for many smaller, non-EVT-capable centers. Even in EVT-capable centers, the ability to screen CTAs for the presence of LVOs can streamline workflow, staffing, and door-to-puncture times by facilitating LVO detection. Machine learning has been employed to automate LVO detection in CTA, which is now in clinical use in a few countries.^7,8^ However, independent external evaluation of these automated LVO detection algorithms have shown only modest sensitivity.^7,8^ Moreover, machine learning algorithms contingent on Hounsfield unit and hemisphere asymmetry information have limited its applicability in patients with bilateral occlusions, such as Moyamoya disease.^9^

In contrast to initial focus on intracranial LVO, recent advancements in neurointerventional devices and cerebrovascular imaging have expanded the application of EVT to medium vessel occlusions. Consequently, the selection of appropriate EVT candidates has become more complex, necessitating advanced imaging. This complexity poses a challenge for less experienced clinicians in making treatment decision.^10^

In the context of current clinical practice, we have developed and validated a fully automated deep learning algorithm to detect comprehensive EVT target vessels. This includes not only the well-known LVOs but also other relevant vessel occlusions. Out development utilized a dataset from multiple centers in Korea, encompassing 2,441 patients with acute ischemic stroke. The deep learning algorithms were specifically designed for 1) selecting the appropriate slices for consistent maximal intensity projection (MIP) image generation, 2) segmenting vessels on MIP images, and 3) identifying vessel occlusions using the vessel segmentation mask. We further validated the algorithm with two independent external datasets.

## Materials and methods

### Datasets

#### Training validation, and internal test

From May 2011 and June 2013, 1,745 patients were recruited from two hospitals. Additionally, 389 patients were included from a university hospital between August 2020 and May 2021. After combining these groups, a total of 2,134 patients who were suspected with ischemic stroke and who underwent CT angiography were initially considered (refer to Supplemental Figure 1). Following the exclusion of 89 patients, the final cohort consisted of 2,045 patients. These were divided into training, validation, and internal test datasets, with 1,277, 144, and 624 patients in each group, respectively.

#### External test

Between April 2011 and July 2013, a total of 71 patient were included from two tertiary hospitals. After excluding 7 patients, 64 were assigned to external test dataset 1. In addition, 337 patients from a university hospital were included for the period between February 2017 and March 2022. After excluding 24 patients from this group, 313 were assigned to external test dataset 2. The study procedure was approved by the institutional review boards of Korea University Guro Hospital (2023GR280), and written consent was waived due to the retrospective and anonymized nature of the study design.

### Definition of Thrombectomy Amenable Vessel Occlusion

In our study, the vascular occlusions we aimed to investigate are termed as Thrombectomy Amenable Vessel Occlusion (TAVO) which include any arterial occlusion involving the intracranial internal carotid artery (ICA), middle cerebral artery (MCA)-M1, and MCA-M2 segments. Intracranial ICA is defined as the segment of the ICA from the petrous part to the MCA-ACA bifurcation. MCA-M1 indicates the MCA segment from the MCA-ACA bifurcation to the MCA branching point, and the MCA-M2 refers to the segment of the MCA ascending vertically along with Sylvian fissure from its branching point. For the subsequent analysis, TAVO patients were divided into intracranial LVO and isolated MCA-M2 occlusion in our study.^11^ In cases of early division of the MCA, a functional rather than traditional angiographic definition was adopted; the short proximal trunk was called the M1 segment and the branches distal to division were defined as M2 segments. To determine the presence of TAVO, each image used in the study was reviewed by an experienced neurologist along with the subject’s MR image (MRI) scans and patient symptom data. The neurologist’s TAVO diagnosis was cross-referenced with the stroke registry, which was independently verified by attending vascular neurologists at each hospital. In case of disagreement, a consensus was made.

### Algorithm Description

#### Slices selection for maximal intensity projection image generation

In order to maintain the uniformity of the input image for the deep learning model, we have developed an automated approach for selecting slices from the source images (Figure 1). These slices are then used to create a maximum intensity projection (MIP) image. We constructed a sagittal bone array by summing the pixel values of the source images. This process automatically outlines the skull, enabling the automatic determination of the vertex and the C1 atlas.^12^ Using this method on the training data, we effectively identified target slices in 1240 cases, which accounts for 97.1% of the total. We calculated the mean and standard deviation of the distance between the vertex and C1 atlas in these patients. For cases falling outside the mean ± 2SD range, we utilized the second method, a deep learning-based algorithm. This model, modified Inception,^13^ classifies CTA source images from the vertex to the C1 atlas as target regions. We used 100 randomly selected cases (4,020 CTA source images), where the first method correctly identified the vertex and C1 atlas. Of these, 89 cases were used for training and 11 cases for validation. The method accurately identified target slices within a 5% margin of error in all cases in the validation dataset. By combining these two techniques, we successfully produced MIP images with a uniform range across all patients in the training and validation dataset. Following skull stripping using our in-house algorithm, we created axial MIP images suitable for the deep learning method, utilizing software developed at our institution.^14^

**Figure 1.**
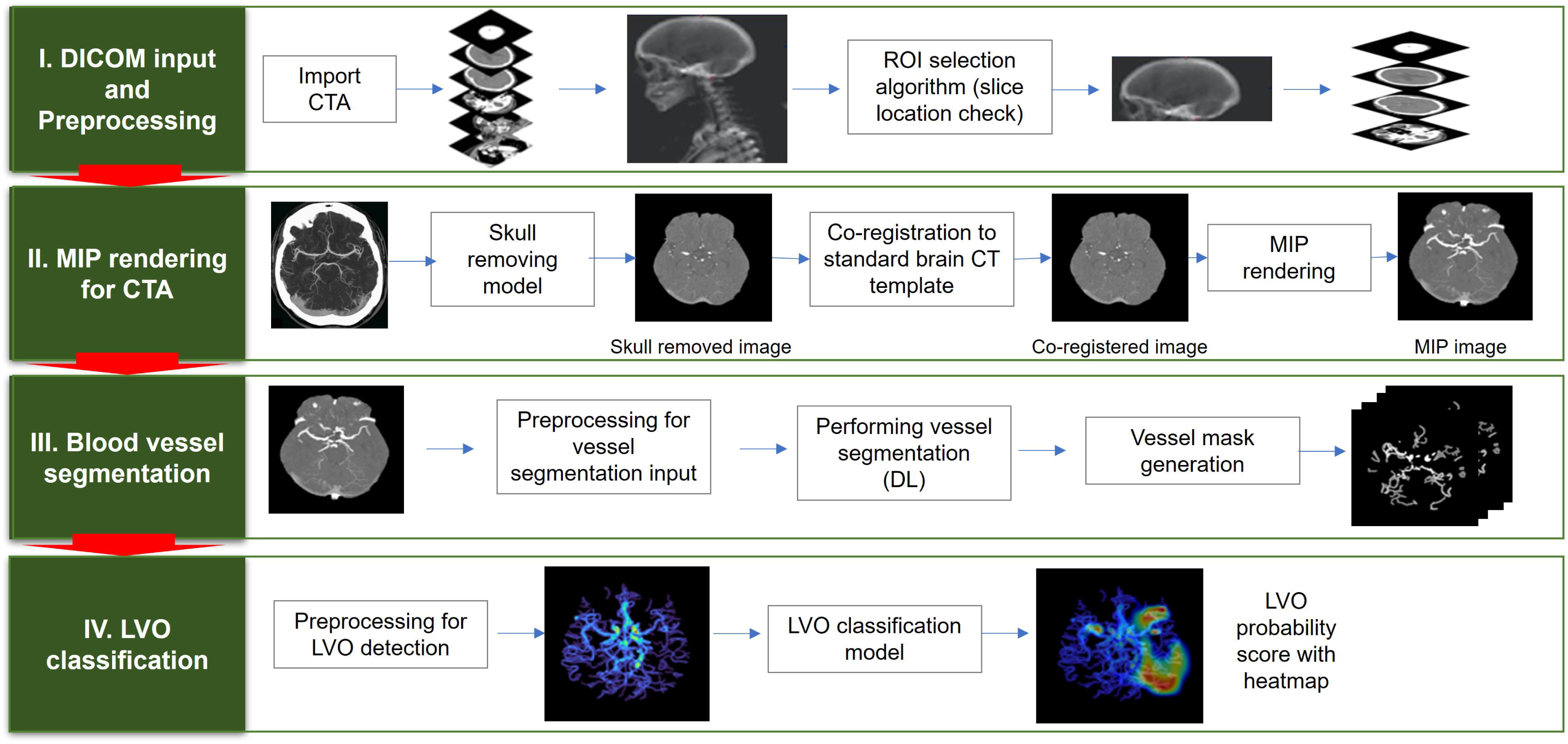
Depiction of the algorithm used to detect automatic Thrombectomy Amenable Vessel Occlusion (TAVO). (I) the acquisition of unprocessed CT angiogram images in DICOM (Digital Imaging and Communications in Medicine) format, accompanied by the automated selection of slices from the vertex and C1 atlas. (II) Skull removal, standard template registration, and maximal intensity projection (MIP) image rendering. (III) Using a rendered axial MIP image to segment blood vessels and merging the blood vessel masks. (IV) Predicting TAVO using the merged blood vessel mask and generating a heatmap that identifies the region that influences the deep learning decision the most.

#### Vessel segmentation on maximal intensity projection image

We developed a 2D U-Net based on the Inception Module specifically for vessel segmentation in axial MIP images.^15^ This model was trained to segment vessels from the generated MIP images. The U-Net architecture integrates structural information from the network with the semantic information from the Inception Module, enabling more precise segmentation of vessels in MIP images. After generating MIP images, researchers manually segmented intracranial arteries in 208 randomly selected patients (16% of whom had TAVO) from the training (n = 189) and validation dataset (n = 19). This manual segmentation was conducted under the supervision of an experienced vascular neurologist (W-S Ryu). The trained model achieved a Dice similarity coefficient of 0.80, indicating strong agreement with the manual segmentation performed by a vascular neurologist.

#### Vessel occlusion detection algorithm

After automatically segmenting blood vessels on the axial MIP image, the vessel masks from each slice were combined to create a two-dimensional compressed image of the vessel masks. These compressed images serve as inputs for our deep learning model, trained for TAVO classification. We observed that using multiple compressed images of vessel masks at constant intervals as input significantly reduced performance compared to using a single compressed image. This was primarily due to overfitting of the algorithm on MCA slices where most TAVOs are located in a specific slice of the 3D volume. The issue of overfitting became more pronounced with an increase in the number of compressed images. Therefore, to mitigate this overfitting, we compressed the segmented vessel masks into a single image for this study. EfficientNetV2 was employed to train the TAVO classification model.^16^ Data augmentation was applied during the training process to prevent overfitting and alleviate domain shift problems. The augmentation algorithm was implemented using albumentations, a Python library for image augmentations.^17^ A batch size of 32 was maintained, with TAVO and non-TAVO cases sampled at a 1:1 ratio in each batch. For the deep learning training, intracranial LVO and isolated MCA-M2 occlusion were combined into a single category as TAVO.

For the training process, we used the AdamW optimizer with a batch size of 32, and a StepLR learning rate Scheduler with a step size of 7 and gamma of 0.1. Additionally, we addressed class imbalance by using a Weighted Random Sampler to sample the TAVO cases more frequently. The training utilized libraries including Python, PyTorch, TensorFlow, Pydicom, OpenCV, ITK, and was performed on an NVIDIA RTX A6000 GPU. The developed software operates on Window 10 or higher.

### Statistical analysis

Data were presented as mean ± standard deviation, median (interquartile range), or number (percentage). To compare baseline characteristics between training and validation, internal test, external test 1, external test 2, we employed ANOVA or Kruskal-Wallis test for continuous variables and chi-square test or Fisher exact test, as appropriate. The diagnostic performance of the algorithm for detecting TAVO was assessed using sensitivity, specificity, positive predictive value (PPV), and negative predictive value (NPV), calculated through receiver operating characteristics (ROC) analysis. The Youden index was utilized to determine the optimal threshold.^18^ We then stratified the patients into two groups: intracranial LVO and isolated MCA-M2 occlusion, and repeated the analysis. In this subgroup analysis, external test 1 and external test 2 dataset were combined due to small number of subjects in the external test 1 dataset. The DeLong test was used to compare the area under the curve (AUC) among the models.^19^ Bootstrap analysis with 1000 repetitions was conducted to calculate 95% confidence intervals for all parameters. All statistical analyses were performed using STATA 16.0 (STATA Corp., Texas, USA), with p < 0.05 considered statistically significant.

## Results

### Baseline characteristics

The mean ages for the training and validation, internal test, external test 1, and external test 2 were 68.7, 68.3, 68.8, and 67.1 years, respectively, as shown in Table 1. The prevalence of male, atrial fibrillation, and history of prior stroke were similar across all groups. However, the occurrence of TAVO was less frequent in the external test 2 dataset compared to the others (13.4% vs. 18.0% to 23.4%). Notably, the CT vendors and imaging parameters, including slick thickness and pixel spacing, significantly varied between the groups (see Supplementary Table 1). For 100 randomly selected cases from the internal test dataset, the mean processing time from the input of source images to the output of results was 178 ± 11 seconds.

**Table 1.** Baseline characteristics of training and validation, internal test, and external test datasets.

|  | Training & validation (n = 1,421) | Internal test (n = 624) | External test 1 (n = 64) | External test 2 (n = 313) | p value |
| --- | --- | --- | --- | --- | --- |
| Age<sup>a</sup> | 68.7 ± 12.6 | 68.3 ± 11.9 | 68.8 ± 12.7 | 67.1 ± 12.3 | 0.45 |
| Sex, men<sup>a</sup> | 797 (56.3%) | 353 (57.0%) | 35 (61.4%) | 199 (63.6%) | 0.11 |
| Onset to image, hour<sup>a</sup> | 20.5 (6.4 – 51.0) | 20.4 (5.3 – 52.9) | 29.5 (16.4 – 72.1) | 22.0 (8.2 – 78.1) | < 0.001<sup>c</sup> |
| Admission NIHSS<sup>a</sup> | 4 (2 – 9) | 4 (2 – 7) | 4 (2 – 7) | 3 (1 – 5) | 0.001<sup>c</sup> |
| Atrial fibrillation<sup>a</sup> | 290 (20.5%) | 116 (18.7%) | 5 (9.0%) | 52 (16.6%) | 0.08 |
| History of prior stroke<sup>a</sup> | 314 (22.2%) | 146 (23.6%) | 14 (24.6%) | 64 (20.5%) | 0.71 |
| Slice thickness of raw image |  |  |  |  | < 0.001 |
| 0.75 mm | 564 (40.0%) | 256 (41.0%) | 59 (92.2%) | 0 |  |
| 1.0 mm | 473 (33.3%) | 232 (37.2%) | 5 (7.8%) | 313 (100%) |  |
| 1.25 mm | 321 (22.6%) | 105 (16.8%) | 0 | 0 |  |
| 1.5 mm | 63 (4.4%) | 31 (5.0%) | 0 | 0 |  |
| Location of TAVO<sup>b</sup> | 255 (18.0%) | 122 (20.0%) | 15 (23.4%) | 42 (13.4%) | 0.08 |
| Any Intracranial ICA | 94 (6.6%) | 26 (4.2%) | 4 (6.3%) | 7 (2.2%) | 0.007 |
| Any M1-MCA | 192 (13.5%) | 84 (13.5%) | 10 (15.6%) | 28 (9.0%) | 0.15 |
| Any M2-MCA | 234 (16.5%) | 101 (16.2%) | 15 (24.4%) | 39 (12.5%) | 0.12 |
| Isolated M2-MCA | 55 (3.9%) | 33 (5.3%) | 5 (7.8%) | 14 (4.5%) | 0.28 |

<sup>a</sup>Clinical data were missing in 4, 5, and 7 patients in training & validation, internal test, and external test 1 datasets, respectively.
<sup>b</sup>Isolated M2-MCA occlusion was considered as TAVO.
<sup>c</sup>Kruskal-Wallis test was used.
MCA indicates middle cerebral artery; NIHSS, National Institutes of Health Stroke Scale; TAVO, Thrombectomy Amenable Vessel Occlusion

### Diagnostic performance for overall TAVO

Representative examples of TAVO detection using deep learning in four patients with intracranial LVO or isolated MCA-M2 occlusion are illustrated in Figure 2. In the internal test dataset, the deep learning algorithm achieved an area under the AUC of 0.950 (95% CI, 0.915 – 0.971, see Figure 3). For the external test datasets 1 and 2, the AUCs were 0.970 (0.897 – 0.997) and 0.971 (0.924 – 0.990), respectively. Using a cutoff threshold of 0.5, the sensitivity ranged from 0.800 to 0.860 (Table 2), and specificity varied from 0.956 to 1.000. The result of the combined external test dataset is visualized in Supplementary Figure 2. Density plots of TAVO probability demonstrated that the deep learning algorithm could robustly differentiate between TAVO and non-TAVO in both internal and external datasets (as shown in Supplementary Figure 3A and 3B).

**Figure 2.**
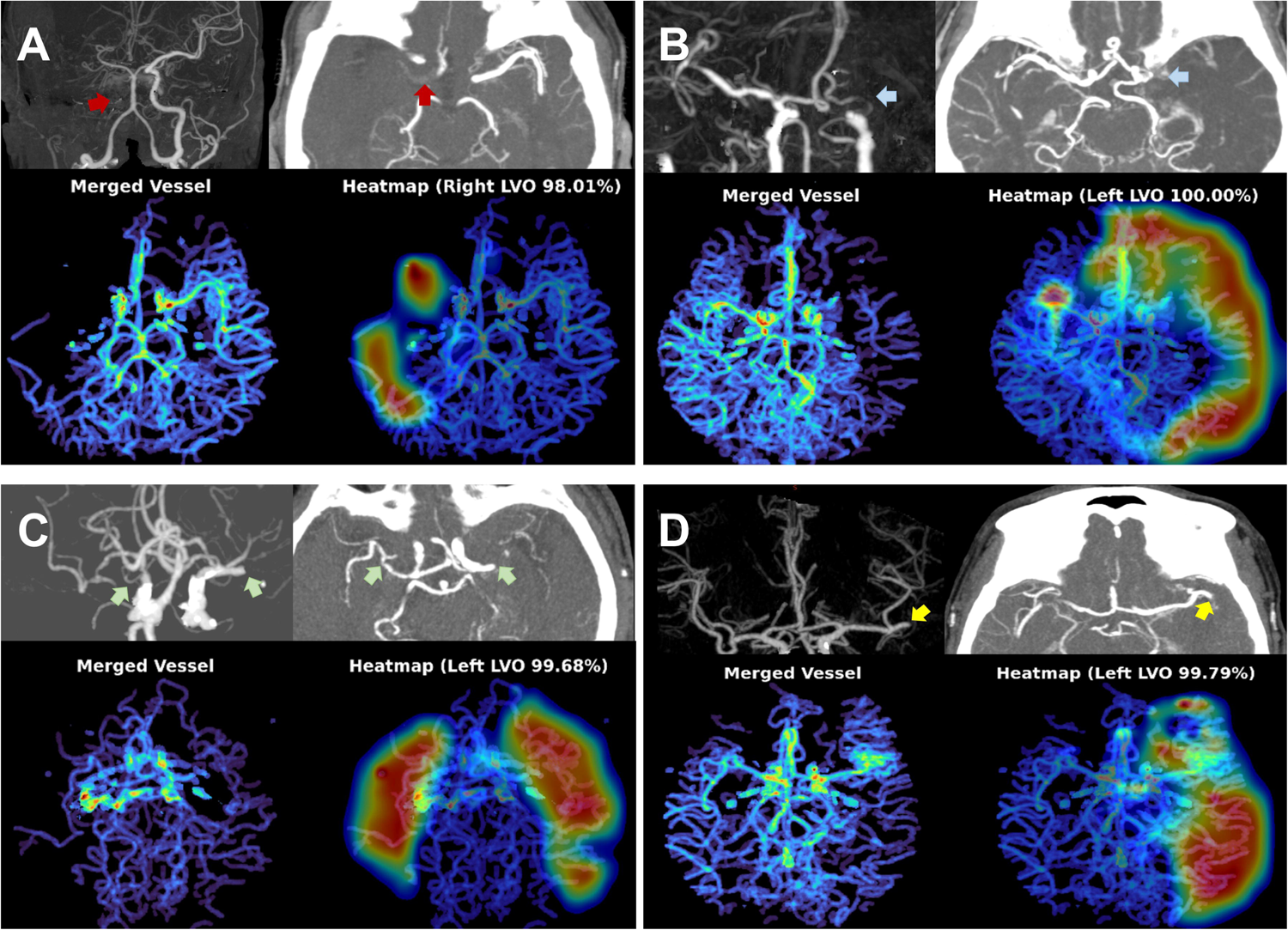
Representative cases for deep learning based Thrombectomy Amenable Vessel Occlusion (TAVO) detection. A, A woman in 70s with cervical internal carotid artery (ICA) occlusion (red arrows) without reconstruction of distal flow. B, A man in 50s man with left distal carotid and left proximal middle cerebral artery (MCA)-M1 occlusion (blue arrows). C, A woman in 70s with bilateral proximal MCA-M1 occlusion (green arrows). D, A woman in 60s with occlusion of the proximal left inferior M2 division (yellow arrows). In all cases, the heatmap visualize the occlusion site or a paucity of distal flow.

**Figure 3.**
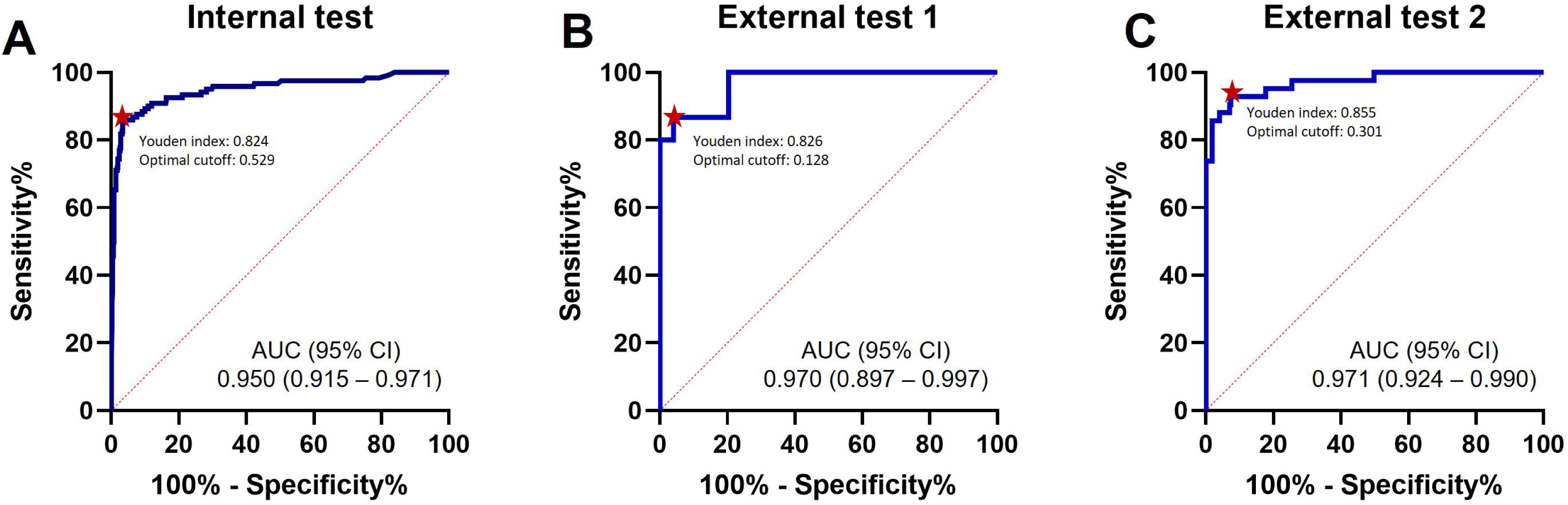
ROC analysis. ROC curves for detection of Thrombectomy Amenable Vessel Occlusion (TAVO) occlusions. (A) Internal test, (B) External test 1, (C) External test 2. Red dots indicate optimal cutoff points with the maximum Youden index. ROC=receiver operating characteristics; AUC=area under the curve.

**Table 2.**
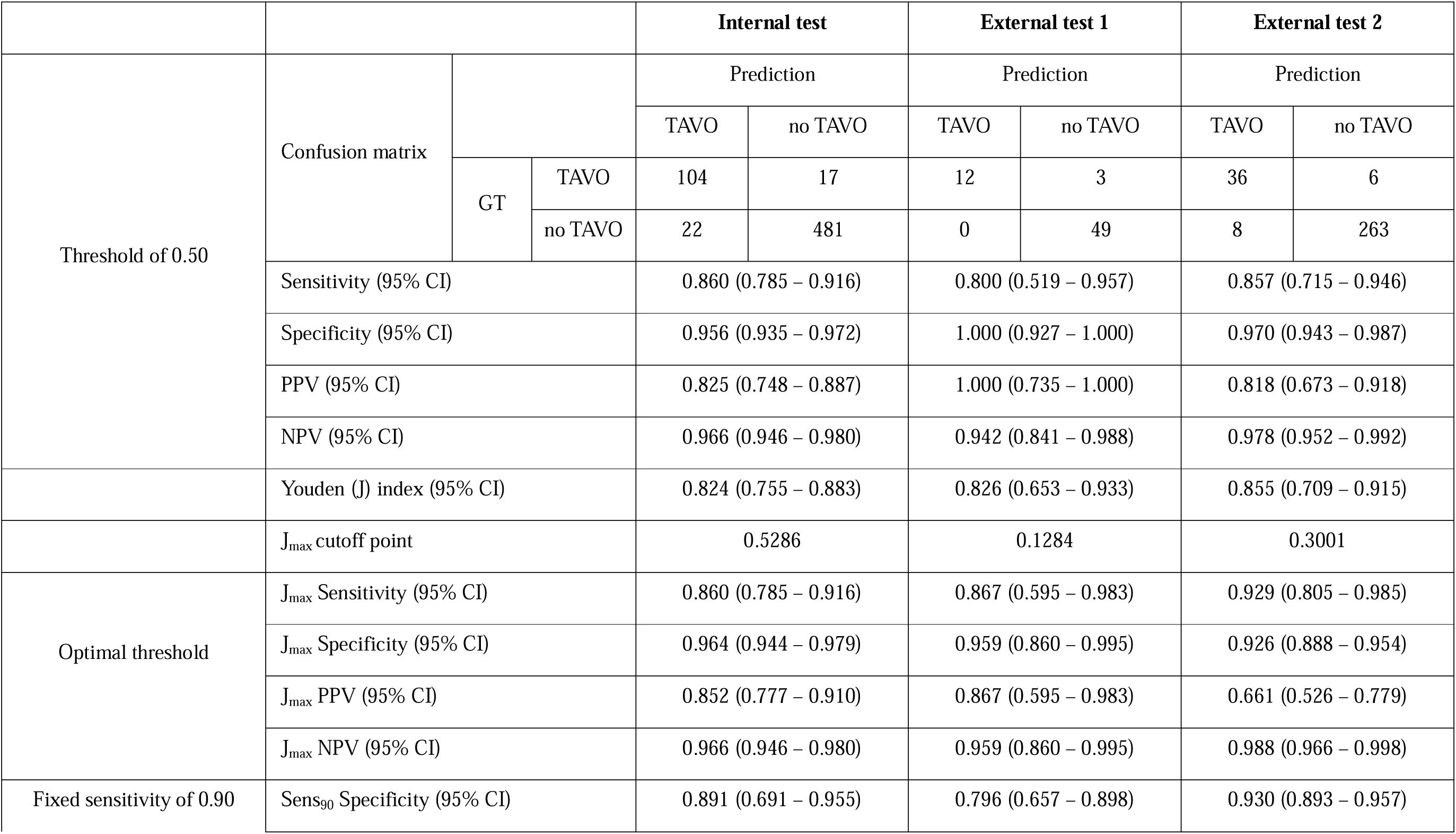
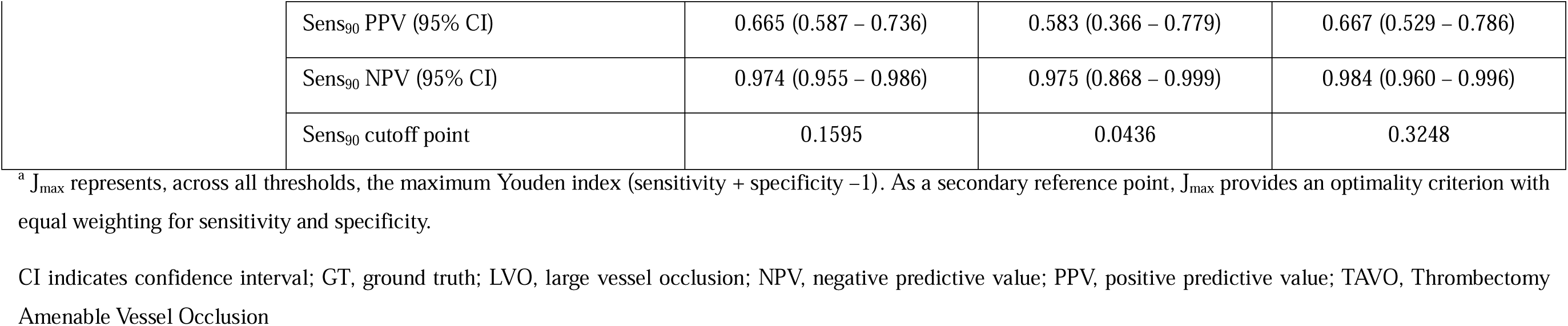
Diagnostic performance of deep learning algorithm detecting TAVO.

The Youden indices in the internal test, external test 1, and external test datasets were 0.824, 0.826, and 0.855, respectively, as detailed in Table 2. In addition, the Youden indices remained stable across a wide range of cutoff points (Supplementary Figure 3C and 3D). At the optimal cutoff points, the sensitivities recorded were 0.860, 0.867, and 0.929, respectively, with corresponding specificities of 0.964, 0.959, and 0.926. When the external test datasets were combined, at the optimal cutoff point, sensitivity, specificity, PPV, and NPV were 0.895 (0.785 – 0.960), 0.934 (0.901 – 0.959), 0.708 (0.589 – 0.810), and 0.980 (0.958 – 0.993), respectively (see Supplementary Table 2). With a fixed sensitivity of 0.900, the specificities and PPVs for the internal test, external test 1, and external test 2 were 0.891, 0.796, and 0.930 and 0.665, 0.583, and 0.667, respectively.

### Diagnostic performance for intracranial Large Vessel Occlusion

Excluding subjects with isolated MCA-M2 occlusion from the analysis significantly improved the performance of the deep learning algorithms. The AUC increased up to 0.967 in the internal dataset and to 0.993 in the combined external datasets, as shown in Table 3 and Supplementary Figure 4. With a cutoff point of 0.5, the sensitivity and specificity in the internal test were 0.943 (0.872 – 0.981) and 0.956 (0.935 – 0.972), respectively. The corresponding values in the combined external dataset were 0.947 (0.823 – 0.994) and 0.975 (0.951 – 0.989).

**Table 3.**
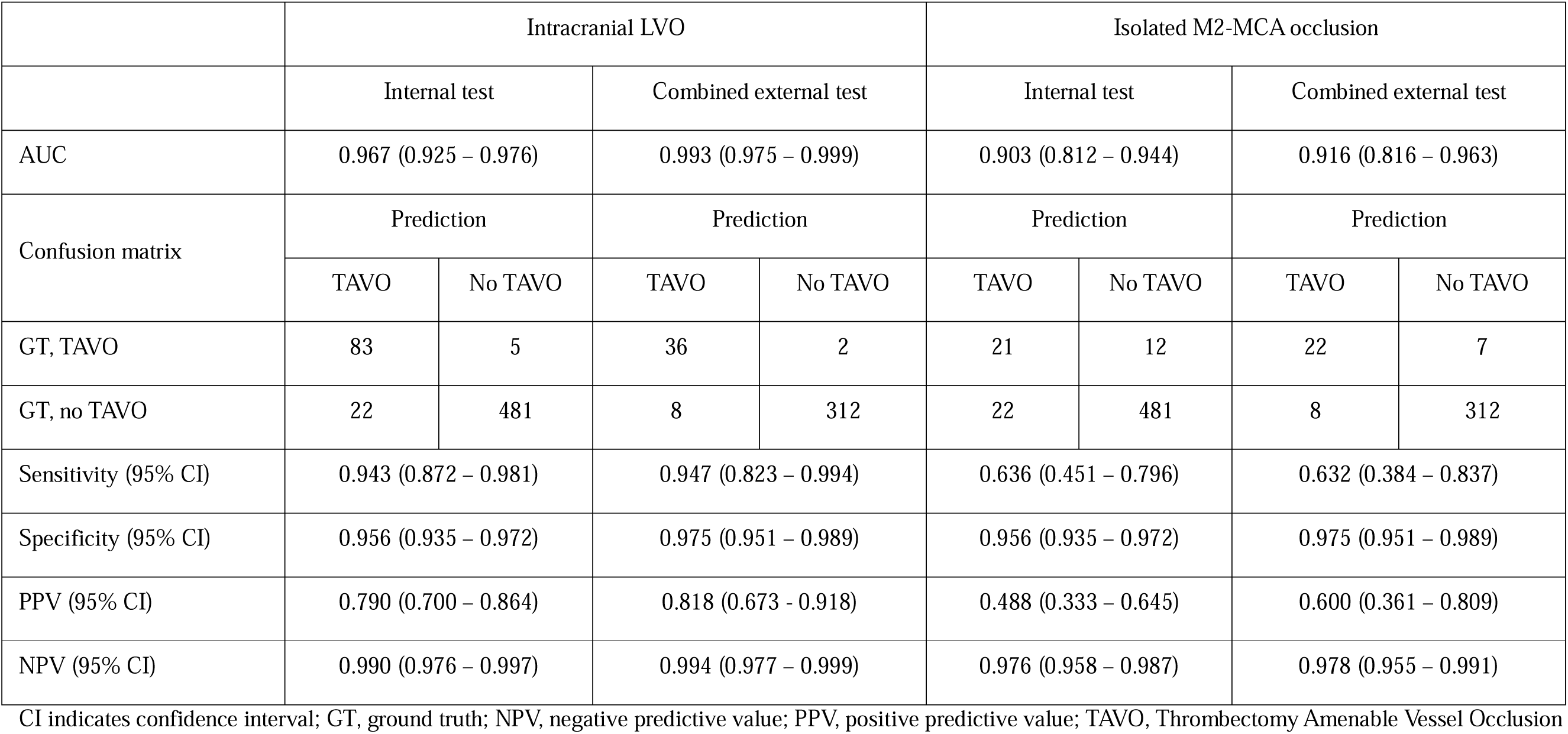
Diagnostic performance of deep learning algorithm stratified by location of occlusion.

### Diagnostic performance for isolated MCA-M2 occlusion

When subjects with intracranial LVO were excluded from the analysis, the diagnostic performance of deep learning algorithm was somewhat diminished. The AUC of internal and combined external datasets were 0.903 (0.812 – 0.944) and 0.916 (0.816 – 0.963), respectively. The sensitivities were 0.636 in the internal test and 0.632 in the combined external dataset. However, the specificities remained high at 0.956 and 0.975, respectively.

### False positive and false negative samples in external test datasets

In the combined external test datasets, eight cases were incorrectly classified as TAVO, as (Supplementary Table 3). Among these, three patients had occlusions in the MCA-M3 or MCA-M4 segments, which are not considered candidate for EVT. Of the nine patients misclassified as non-TAVO, seven had isolated occlusions in the MCA-M2 segment. Two patients with false negative results and intracranial LVO had marginal TAVO probability score of 0.155 and 0.355, respectively.

## Discussion

In this study, we developed a fully automated deep learning algorithm to detect intracranial anterior circulation arterial occlusions in CTA, which are likely candidates for EVT in hyperacute ischemic stroke. The algorithm underwent external validation in two different datasets and demonstrated high diagnostic sensitivity and specificity. When analyzing occlusion sites separately, the algorithm performed well for intracranial LVO. Although the performance for isolated MCA-M2 occlusion was slightly lower than for intracranial LVO, it remained competitive.

Until recently, a number of studies have reported on AI detection of LVO or TAVO (see Supplementary Table 4). However, these algorithms were either lacked external validation in previous research, or if validated, it was in a limited number of cases.^20–23^ Moreover, earlier studies reported that the artificial intelligence algorithms achieved AUC scores ranging from 0.74 to 0.86, which may not sufficiently support less experienced physicians.^20–23^ Notably, in external validation sets with adequate sample sizes, our deep learning algorithm achieved AUCs of 0.961, 0.993, and 0.913 for total TAVO, intracranial LVO, and isolated MCA-M2 occlusion, respectively.

The ability of our deep learning algorithm to accurately predict isolated MCA-M2 occlusion with good performance is of significant importance. Although isolated MCA-M2 occlusion is an important and emerging target of the EVT,^24^ it was reported that the rate of misdiagnosis for isolated MCA-M2 occlusion is substantially higher than that for intracranial LVO even among the neuroimaging specialists (35.0% vs. 9.7%, respectively),^25^ and the use of previously developed deep learning models has yielded worse results (50.8%).^20^ This characteristic of isolated MCA-M2 occlusion have influenced our model as well, leading to improved specificity and NPV compared to previous studies,^20,21^ despite with somewhat less satisfactory sensitivity and PPV. This issue was largely due to false negatives in cases of short-segment (where collaterals reconstituted the M2 segment immediately distal to the occlusion) and incomplete (with antegrade flow) occlusions, where the reduction in the inter-hemispheric vessel density was too small for the algorithm to detect. However, in terms of TAVO detection, these occlusions missed due to false negative may not be ideal candidates for emergent intra-arterial thrombectomy.^26^

Our study may have a few potential clinical implications. First, our deep learning model can provide decent assistance to healthcare professionals who may not have much experience with ischemic strokes. Regardless of the reason for admission, the majority of ischemic strokes are initially encountered by non-specialized physicians.^27^ In such scenarios, identifying vascular occlusion and its location in brain imaging is a complex and demanding task, potentially delaying appropriate stroke treatment. Second, considering that our model processes vascular images in less than 180 seconds, it could significantly reduce the time taken to make EVT decision. In clinical practice, formal interpretation of brain imaging often requires several hours, and sometimes even more than a day.^28^ Therefore, rapid primary interpretation of vascular status through our model can play a crucial role in shortening the time to initiate EVT. Third, the objective and reproducible nature of our artificial intelligence software can aid in refining EVT process. This includes the preparation protocol for interventional devices based on occlusion patterns, which is another important factor in reducing reperfusion time.

### Limitation of the study

One limitation of our study is the relatively small dataset size used for training the deep learning algorithm. This is particularly true for isolated MCA-M2 occlusion, where fewer cases were enrolled compared to intracranial LVO, influencing the sensitivity and PPV performance of the model. The second caveat is that this study was conducted within a single country and given the higher prevalence of intracranial atherosclerosis in East Asian populations compared to Western ones, the assessment of the complete occlusion over pre-existing stenosis may have been challenging.^29^ Such factors could contribute to model’s less favorable performance, underscoring the need for further research involving diverse ethnic groups. Therefore, radiologists and neurologists should be aware of the potential and causes of false negatives in the algorithm’s output.

### Future Directions

Collateral score and collateral status on CTA serve as important tools in evaluating collateral circulation in patients with TAVO.^26^ In CTA in patients with TAVO, collateral score and collateral status are used as important parameters to evaluate cerebral hemodynamics and tissue perfusion. These assessments help understand the adequacy of compensatory collateral circulation, which is essential for determining treatment strategies and predicting tissue viability in patients with ischemic stroke. Our developed system can estimate the course of collateral circulation by detecting and displaying the location of TAVO as a heat map. Collateral status involves a comprehensive assessment of the collateral circulation, considering factors such as collateral filling rate, extent of collateral vessels, and final tissue perfusion achieved through collateral flow.^30^ This assessment plays a pivotal role in predicting the likelihood of ischemic tissue salvage. Appropriate adjuvants contribute to increasing the likelihood of viable tissue despite vascular occlusion, potentially influencing treatment decisions such as endovascular reperfusion therapy. Therefore, auxiliary evaluation of collateral status may be considered in future studies.

## Conclusion

We developed and validated a novel, fully automated deep learning algorithm derived from CTA to detect vascular occlusion suitable for EVT. While the algorithm could benefit from further improvements and real-world clinical evaluations, its potential as a tool to assist in the diagnosis of acute ischemic stroke in patients through detection of TAVO has been firmly established.

## Data Availability

All data produced in the present study are available upon reasonable request to the authors.

