## Supplementary Material for "Automated Identification of Thrombectomy Amenable Vessel Occlusion on Computed Tomography Angiography using Deep Learning"

Jung Hoon Han et al.

Supplementary Table 1. Comparison of imaging parameters among training & validation, internal test, external test datasets

| | Training & validation (n = 1,421) | Internal test (n = 624) | External test 1 (n = 64) | External test 2 (n = 313) | p value |
|---|---|---|---|---|---|
| **Vendor** | | | | | < 0.001 |
| GE | 321 (22.6%) | 131 (21.0%) | 0 | 0 | |
| Philips | 4 (0.3%) | 0 | 5 (7.8%) | 0 | |
| Siemens | 1,096 (77.1%) | 493 (79.0%) | 59 (92.2%) | 290 (92.7%) | |
| Toshiba | 0 | 0 | 0 | 23 (7.4%) | |
| **Peak Kilovoltage, kVp** | | | | | < 0.001 |
| 80 | 1 (0.1%) | 0 | 4 (6.3%) | 0 | |
| 90 | 0 | 0 | 0 | 41 (13.1%) | |
| 100 | 113 (8.0%) | 103 (16.5%) | 55 (85.9%) | 251 (80.2%) | |
| 120 | 1,294 (91.1%) | 521 (83.5%) | 5 (7.8%) | 21 (6.7%) | |
| 140 | 13 (0.9%) | 0 | 0 | 0 | |
| **Data collection diameter, mm** | | | | | < 0.001 |
| 240 | 0 | 0 | 0 | 5 (1.6%) | |
| 320 | 321 (22.6%) | 131 (21.0%) | 0 | 16 (5.1%) | |
| 400 | 0 | 0 | 0 | 2 (0.6%) | |
| 500 | 1,100 (77.4%) | 493 (79.0%) | 64 (100%) | 290 (92.7%) | |
| **Tube current, mA** | | | | | < 0.001 |
| 101~319 | 260 (18.3%) | 119 (19.1%) | 0 | 98 (31.3%) | |
| 320 | 459 (32.3%) | 187 (30.0%) | 0 | 0 | |
| 321~995 | 702 (49.4%) | 318 (50.1%) | 64 (100%) | 215 (68.7%) | |
| **Pixel spacing, mm** | | | | | < 0.001 |
| 0.30 ~ 0.38 | 451 (31.7%) | 192 (30.8%) | 10 (15.6%) | 3 (1.0%) | |
| 0.39 ~ 0.40 | 676 (47.6%) | 340 (54.5%) | 16 (25.0%) | 16 (5.1%) | |
| 0.41 ~ 0.78 | 294 (20.7%) | 92 (14.7%) | 38 (59.4%) | 294 (93.9%) | |

Supplementary Table 2. Diagnostic performance of deep learning algorithm detecting intracranial LVO + MCA-M2 occlusion in combined external dataset

| | | | |
|---|---|---|---|
| Threshold of 0.50 | Confusion matrix | Prediction | |
| | | LVO | No LVO |
| | GT, LVO | 48 | 9 |
| | GT, no LVO | 8 | 312 |
| | Sensitivity (95% CI) | 0.842 (0.721 – 0.925) | |
| | Specificity (95% CI) | 0.975 (0.951 – 0.989) | |
| | PPV (95% CI) | 0.857 (0.738 – 0.936) | |
| | NPV (95% CI) | 0.972 (0.947 – 0.987) | |
| | Youden (J) index (95% CI) | 0.829 (0.712 – 0.892) | |
| | $J_{max}$ cutoff point | 0.3008 | |
| Optimal threshold | $J_{max}$ Sensitivity (95% CI) | 0.895 (0.785 – 0.960) | |
| | $J_{max}$ Specificity (95% CI) | 0.934 (0.901 – 0.959) | |
| | $J_{max}$ PPV (95% CI) | 0.708 (0.589 – 0.810) | |
| | $J_{max}$ NPV (95% CI) | 0.980 (0.958 – 0.993) | |
| Fixed sensitivity of 0.90 | $Sens_{90}$ Specificity (95% CI) | 0.863 (0.807 – 0.971) | |
| | $Sens_{90}$ PPV (95% CI) | 0.542 (0.437 – 0.644) | |
| | $Sens_{90}$ NPV (95% CI) | 0.982 (0.959 – 0.994) | |
| | $Sens_{90}$ cutoff point | 0.1776 | |

GT indicates ground truth; NPV, negative predictive value; PPV, positive predictive value; TAVO, Thrombectomy Amenable Vessel Occlusion

Supplementary Table 3. Review of false positive and false negative cases in the external test dataset

| False positive | | | | | | | |
|---|---|---|---|---|---|---|---|
| | Age | Sex | LVO probability score | Stroke subtype | Initial NIHSS score | Location of vessel occlusion | Revascularization therapy |
| case 1 | 60s | male | 0.8883 | 1 | NA | Right MCA-M3 | Yes |
| case 2 | 50s | female | 0.8832 | 2 | NA | No occlusion | No |
| case 3 | 60s | male | 0.9138 | 1 | 5 | No occlusion | No |
| case 4 | 60s | male | 0.7683 | 2 | 1 | Left MCA-M3 | No |
| case 5 | 80s | female | 0.5188 | 1 | 1 | No occlusion | No |
| case 6 | 60s | female | 0.5044 | 3 | 0 | No occlusion | No |
| case 7 | 80s | female | 0.8909 | 6 | 5 | Right M3 or M4 | No |
| case 8 | 70s | male | 0.5021 | 3 | 7 | No occlusion | No |
| False negative | | | | | | | |
| | Age | Sex | LVO probability score | Stroke subtype | Initial NIHSS score | Location of vessel occlusion | Revascularization therapy |
| case 9 | 70s | male | 0.0782 | 1 | 7 | Left MCA-M2 | No |
| case 10 | 50s | female | 0.4194 | 6 | 11 | Left MCA-M2 | Yes |
| case 11 | 60s | male | 0.304 | 1 | 1 | Right MCA-M2 | No |
| case 12 | 80s | male | 0.1551 | 1 | 3 | Right MCA-M1 | No |
| case 13 | 50s | female | 0.0125 | 3 | 13 | Left MCA-M2 | Yes |
| case 14 | 50s | male | 0.355 | 1 | 2 | Left MCA-M1 | No |
| case 15 | 70s | female | 0.1799 | 1 | 2 | Right MCA-M2 | No |
| case 16 | 70s | female | 0.0424 | 7 | 8 | Right MCA-M2 | No |
| case 17 | 40s | male | 0.0447 | 2 | 8 | Right MCA-M2 | No |

MCA indicates middle cerebral artery; NIHSS, National Institutes of Health Stroke Scale; TAVO, Thrombectomy Amenable Vessel Occlusion

Supplementary Table4. Summary of related previous studies.

| Author | End point | n | Models | Results | Modality | Remark | Year |
|---|---|---|---|---|---|---|---|
| Olive-Gadea et al.(20) | LVO prediction | 1453 | Densenet161 | ·AUC (0.87)<br>·Sen (0.83)<br>·Spec(0.71) | NCCT | Validation of prior ML algorithm | 2020 |
| Meng et al.(21) | ·LVO classification<br>·Clinical outcome prediction | 8650 | Inception-V1 | ·AUC (0.74)<br>·Sen (0.61)<br>·Spec (0. 74) | CTA | Validation of prior ML algorithm | 2022 |
| Czap et al.(22) | LVO classification | ·Training - 870<br>·Internal validation- 441 | DeepSymNet-v2 | ·AUC (0.80) | CTA (MSU) | Small external validation test size (n=68) | 2022 |
| Matsoukas et al. (23) | LVO classification | 1822 | Viz LVO | ·AUC (0.86)<br>·Sen (74.6)<br>·Spec (91.1) | CTA | Validation of prior ML algorithm | 2022 |
| This study | LVO classification | ·Training - 1422<br>·Internal validation - 629<br>External validation - 390 | ·Vessel Segmentation - U-Net<br>·LVO Classification - EfficientNetV2 | ·AUC (0.96)<br>·Sen (0.80)<br>·Spec (0.97) | CTA | Novel development<br>Two external validation set | Present |

AUC indicates area under the curve; CTA, computed tomography angiography; ML, machine learning; MSU, mobile stroke unit; NCCT, non- contrast computed tomography; TAVO, Thrombectomy Amenable Vessel Occlusion

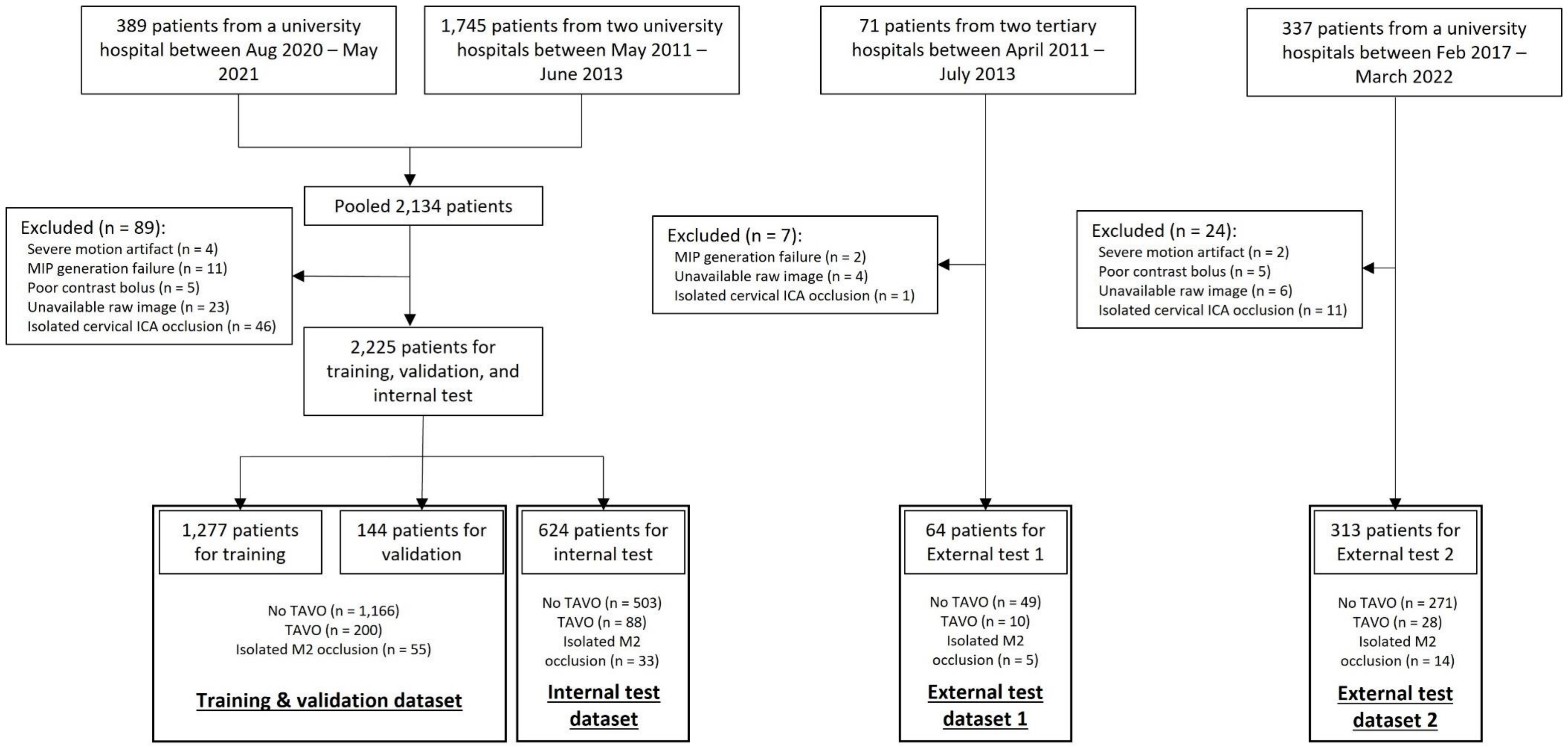


Supplementary Figure 1. Study flow chart. TAVO indicates Thrombectomy Amenable Vessel Occlusion.

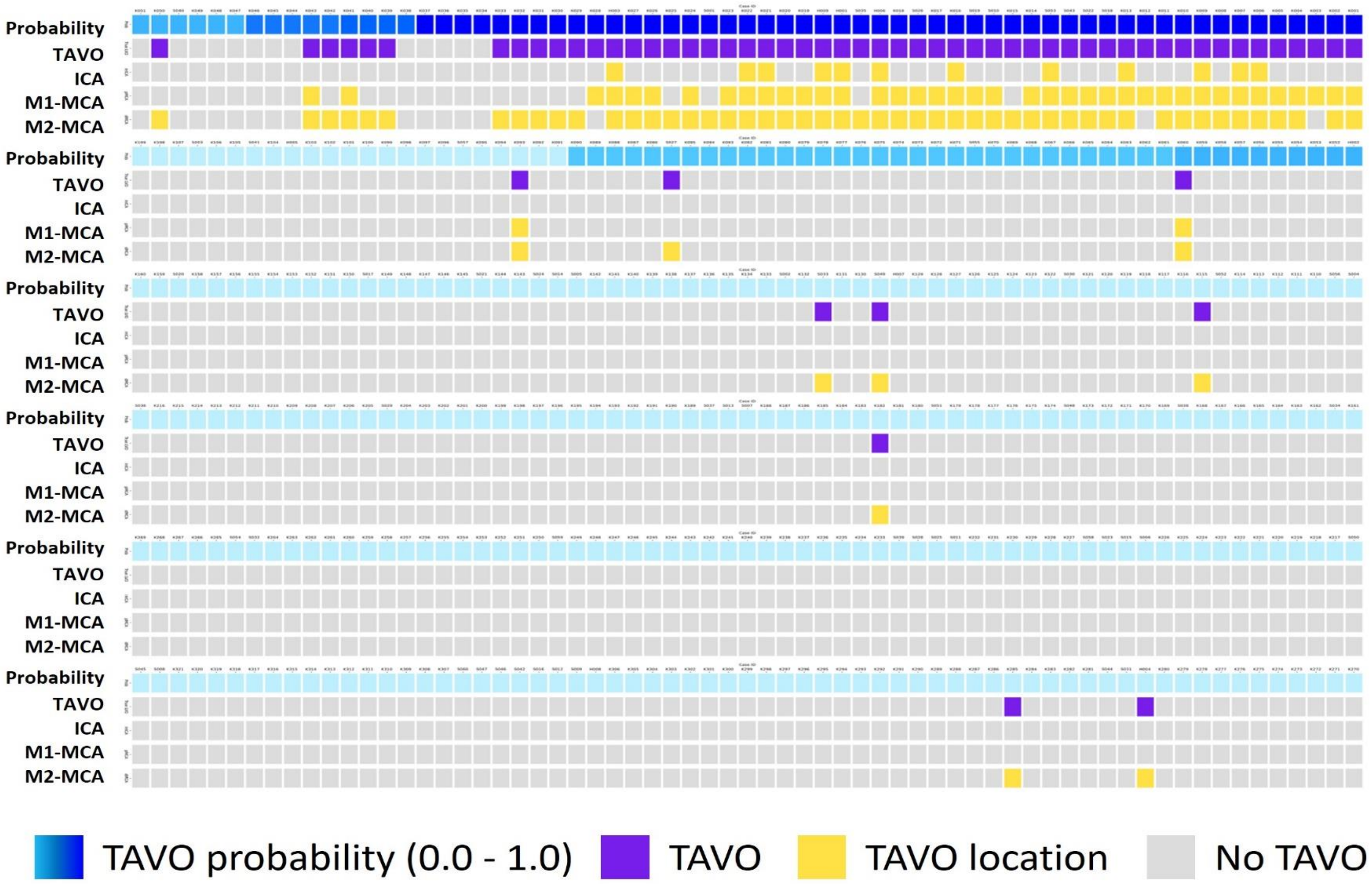


Supplementary Figure 2. Probability of Thrombectomy Amenable Vessel Occlusion (TAVO), location of TAVO, and ground truth label for TAVO for each subject in the combined external test dataset. Each column indicates each subject. The first row indicates TAVO probability score. The second row indicates the ground truth label of TAVO. The third, fourth, and fifth rows indicate the location of TAVO. ICA indicates internal carotid artery; MCA, middle cerebral artery

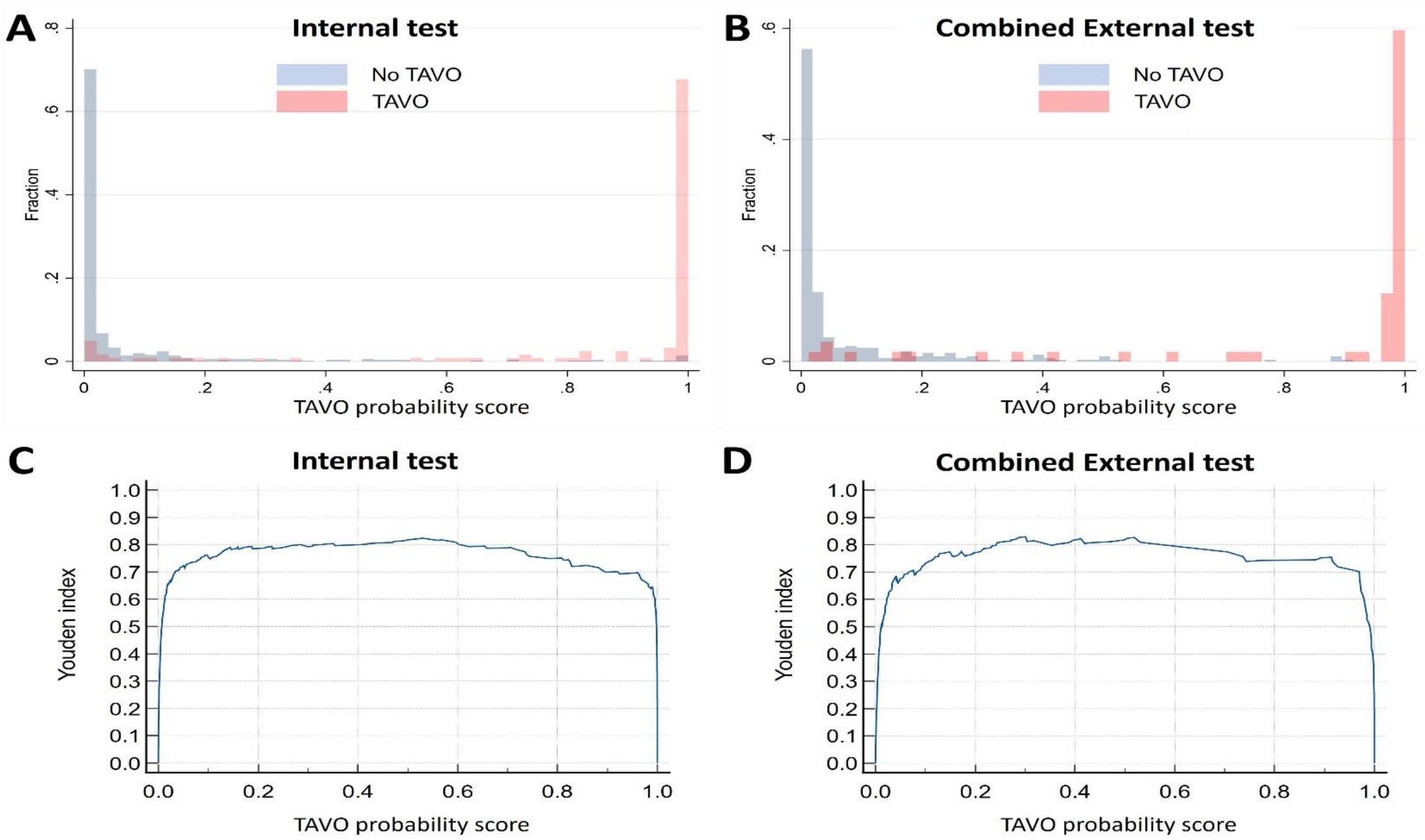


Supplementary Figure 3. Probability density in (A) internal test dataset and (B) combined external test dataset. Change of Youden index by probability cutoff in (C)internal test dataset and (D) combined external test dataset.

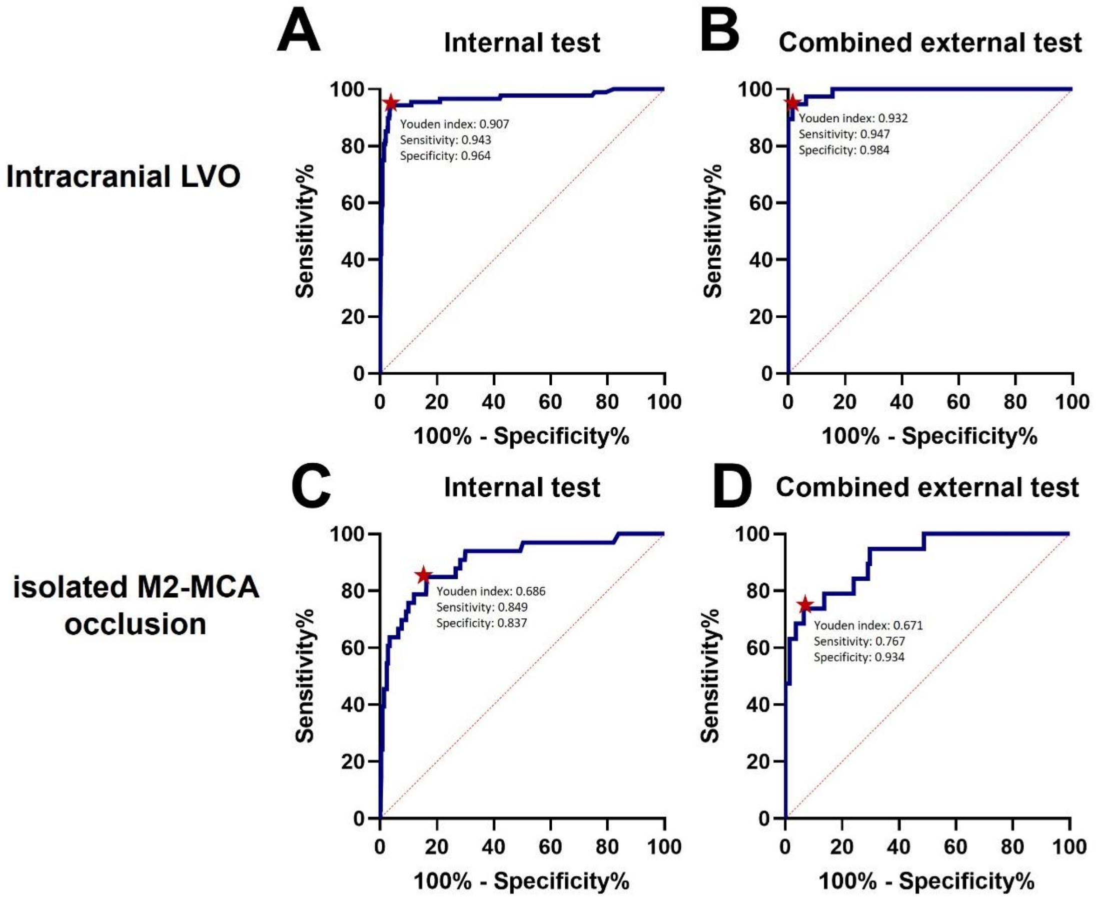


Supplementary Figure 4. ROC analysis. ROC curves for detection of intracranial LVOs in (A) Internal test and (B) Combined external dataset. ROC curves for detection of isolated M2-MCA occlusion in (C) Internal test and (D) Combined external datasets. Red dots indicate optimal cutoff points with the maximum Youden index. AUC indicates area under the curve; LVO, large vessel occlusion; ROC, receiver operating characteristics.
